# Longitudinal Multimodality Imaging of Cardiovascular Abnormalities Following Definitive Chemoradiation for Lung Cancer: A Retrospective Cohort Study

**DOI:** 10.64898/2026.11.19.26367918

**Authors:** Troy Nguyen, Vivian Salama, Joseph A. Schmidlen, Peyton M. Lilly, Sijin Wen, Nathaniel Dunham, R Alfredo Siochi, Irfan Zeb, Bavly Naklah, David A. Clump, Phillip M. Pifer, Mina F. Hanna

**Author notes:** **Corresponding authors:** Mina F. Hanna, MD, MSc, Associate Professor of Radiology, Section Chief, Cardiovascular and Thoracic Imaging Vice Chair of Faculty Development, Department of Radiology, West Virginia University School of Medicine, Morgantown, WV, USA PO Box 9235, 64 Medical Center Drive Morgantown, WV 26506, Vivian Salama, MD, PhD, MSc, Research Assistant Professor, Departments of Medical Oncology, Radiation Oncology and Radiology West Virginia University School of Medicine, WVU Cancer Institute PO Box 9162, 1 Medical Center Drive 1838 Cancer Institute, Morgantown, WV 26506-9162. These authors contributed equally and shared the first authorship (Troy Nguyen, Vivian Salama). **Institution from which the work originated:** Department of Radiology, West Virginia University School of Medicine 64 Medical Center Drive, Morgantown, WV 26506, USA.

## Abstract

**Background/Objectives:** Cardiovascular complications following definitive chemoradiation therapy (CRT) for lung cancer are well recognized, but their longitudinal imaging manifestations remain incompletely characterized. We aimed to characterize cardiovascular abnormalities across multiple imaging modalities and electrocardiography during lung cancer survivorship.

**Methods:** This retrospective single-center study included patients treated with definitive CRT between 2013 and 2025 who underwent baseline and follow-up cardiovascular assessments. Findings from computed tomography, echocardiography, myocardial perfusion imaging, coronary angiography, and electrocardiography were categorized into exploratory, literature-informed cardiovascular phenotype domains across five follow-up intervals. Longitudinal patterns were evaluated using generalized estimating equations (GEE). and exploratory associations with clinical cardiovascular adverse events (CVAEs) were assessed using χ² or Fisher’s exact tests.

**Results:** Eighty-six patients were included (mean age, 66 ± 9.1 years; 46 women). Baseline coronary, thoracic aortic, and valvular calcification were common, affecting 72%, 88%, and 50% of patients, respectively. During the first 0–6 months, reduced LV systolic function (53%), valvular dysfunction (35%), and pericardial effusion (19%) were observed. Pericardial effusion was observed in 37% at 1-2 years, while calcification progression and pulmonary vascular abnormalities were identified during later follow-up intervals. Longitudinal analyses suggested positive associations between successive follow-up categories and calcification progression (average OR, 4.30), pulmonary artery enlargement (OR, 1.49; 95% CI, 1.10-2.02; *P = 0.01*), right-heart abnormalities (average OR, 1.65), and arrhythmia (OR, 1.76; 95% CI, 1.30-2.38; *P < 0.001*). Exploratory associations between imaging abnormalities and clinical CVAEs were also observed.

**Conclusions:** Longitudinal multimodality cardiovascular assessments identified heterogeneous cardiovascular abnormalities across follow-up intervals following definitive CRT for lung cancer. Larger prospective studies are needed to validate these observations.

## INTRODUCTION

Definitive concurrent chemoradiation therapy (CRT) is a standard curative-intent treatment for unresectable locally advanced lung cancer (1). Consolidation durvalumab after CRT has improved progression-free and overall survival, increasing the population of longer-term survivors potentially at risk for late cardiovascular complications (2,3). Thoracic CRT may affect multiple cardiovascular structures, including the pericardium, myocardium, valves, coronary arteries, great vessels, pulmonary vasculature, and conduction system, with abnormalities emerging months to years after treatment (4–7).

Multimodality cardiovascular imaging plays a central role in detecting subclinical structural and functional abnormalities before the onset of overt cardiovascular disease, providing opportunities for earlier diagnosis, risk stratification, and clinical management guidance (4,7–9). However, existing evidence has mainly focused on individual cardiovascular abnormalities, single imaging modalities, or clinical endpoints rather than integrated longitudinal phenotype evolution (10–13). Multidisciplinary guidelines recommend risk-adapted surveillance after thoracic radiotherapy but acknowledge limited longitudinal evidence to define optimal strategies (7,14). This gap is particularly relevant to radiologists because routine post-treatment imaging may reveal subclinical cardiovascular phenotypes before or without clinically recognized cardiovascular events.

Furthermore, much of the long-term radiation cardiovascular literature derives from breast cancer and lymphoma cohorts (15,16). Longitudinal multimodality imaging data in lung cancer patients remain limited despite higher cardiac radiation exposure, substantial baseline cardiopulmonary comorbidity, and competing cardiopulmonary risk factors (10,13,17). Appalachian populations are particularly understudied despite disproportionate tobacco-related disease, lung cancer burden, cardiovascular comorbidity, and healthcare disparities (18,19).

Therefore, the aim of this study was to characterize the temporal evolution of cardiovascular phenotypes after definitive CRT for lung cancer using longitudinal multimodality imaging, electrocardiography (ECG), and clinical data.

## MATERIALS & METHODS

### Study design

This HIPAA-compliant retrospective cohort study was approved by the Institutional Review Board, which waived written informed consent. Adults with biopsy-confirmed lung cancer treated with definitive CRT from January 2013 to December 2025 were identified. Clinical data were obtained from the electronic health record, and imaging studies were analyzed from PACS. This cohort overlaps with a prior study evaluating cardiovascular morbidity and exploratory prediction and association analysis using baseline clinical and dosimetric variables (20). The present study evaluates longitudinal multimodality imaging and ECG phenotype evolution across prespecified intervals using repeated-measures analyses.

### Cohort Selection

Eligible patients were adults (>18 years) with biopsy-proven lung cancer who underwent definitive CRT, had complete treatment data, and had baseline and at least one follow-up cardiovascular evaluation. Eligible histology included locally advanced non-small cell lung cancer (NSCLC), limited-stage SCLC, and select oligometastatic NSCLC treated with definitive CRT. Exclusion criteria were palliative or non-definitive treatment, pulmonary metastases from a non-pulmonary primary malignancy, early-stage NSCLC treated with surgery, stereotactic body radiotherapy, hypofractionated radiotherapy, or more than one course of thoracic radiotherapy before a documented cardiovascular adverse event (CVAE). Patients were followed from CRT initiation through last imaging, last clinical encounter, or death. Of 386 screened patients, 86 met eligibility criteria (Figure 1).

**Figure 1.**
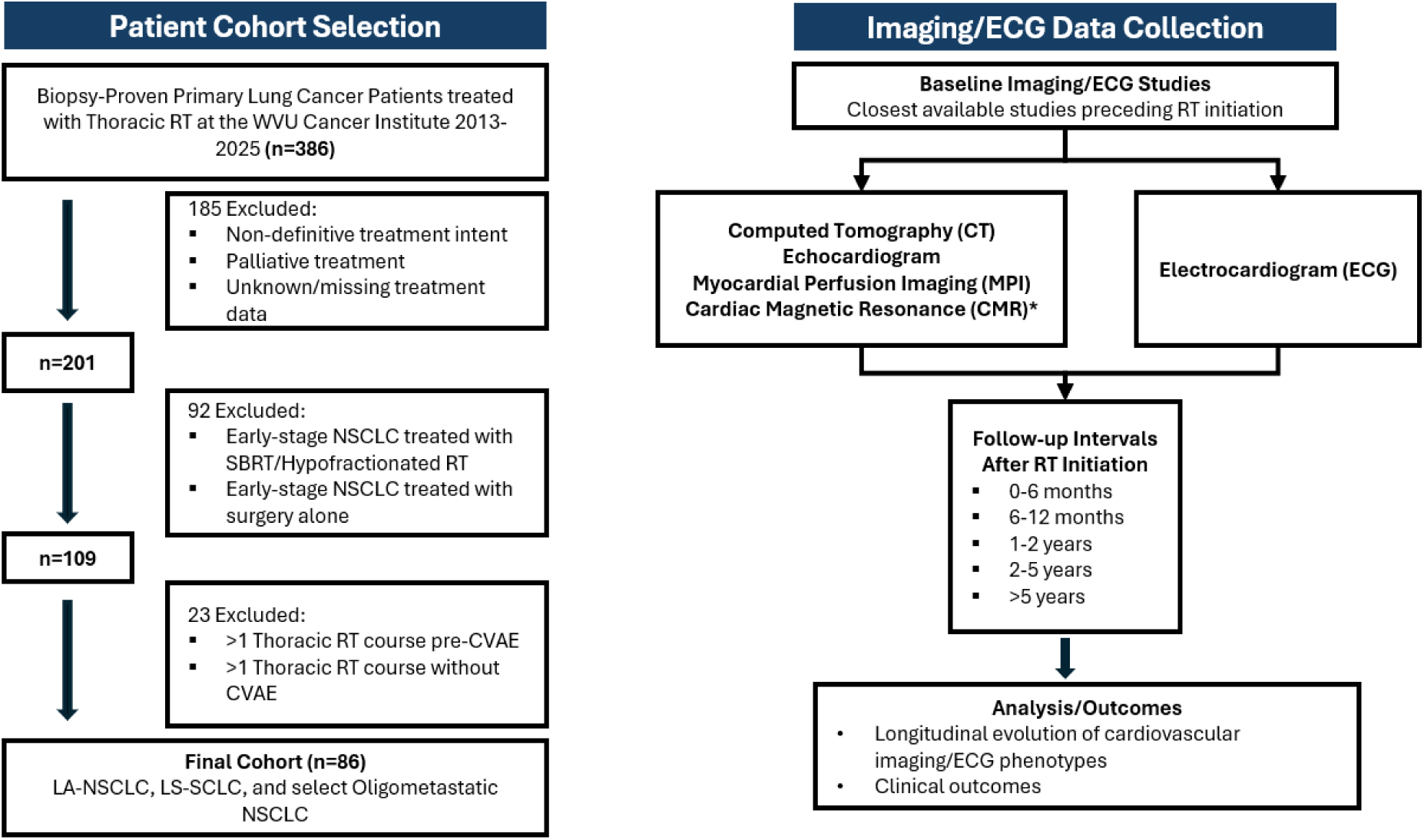
Study cohort selection and longitudinal multimodality cardiovascular imaging/ECG data collection. Flow diagram shows initial patient screening, exclusion criteria, final cohort selection, and collection of multimodality imaging and ECG data across baseline and five follow-up intervals after definitive CRT. *Cardiac MRI data were insufficient for analysis. Abbreviations: CMR = cardiac magnetic resonance; CRT = chemoradiation therapy; CVAE = cardiovascular adverse event; ECG = electrocardiography; LA-NSCLC = locally advanced non-small cell lung cancer; LS-SCLC = limited-stage small cell lung cancer; MPI = myocardial perfusion imaging; NSCLC = non-small cell lung cancer; RT = radiotherapy; SBRT = stereotactic body radiation therapy

Baseline imaging and ECG were defined as the closest examination preceding CRT initiation. Follow-up studies were categorized into 0-6 months, 6-12 months, 1-2 years, 2-5 years, and >5 years after CRT initiation.

### Imaging acquisition and review

Cardiovascular imaging studies performed as part of routine care were reviewed from electronic health records and PACS, including chest CT, echocardiography, myocardial perfusion imaging (MPI), and invasive coronary angiography when available. Cardiac MRI data were insufficient for longitudinal analysis.

Pre-CRT imaging established baseline status. Post-CRT studies were reviewed across follow-up intervals with access to prior examinations for direct comparison. CT images were reviewed by a diagnostic radiology resident under supervision of a fellowship-trained cardiothoracic radiologist, with uncertain cases adjudicated by consensus. Echocardiographic, MPI, coronary angiographic, and ECG findings were abstracted from finalized reports.

At each interval, imaging and ECG findings were classified relative to baseline as stable, improved, new, or worsened. When multiple examinations were available within an interval, the most severe new or worsened abnormality was recorded. Standardized variable definitions adapted from published imaging literature were used throughout review, particularly for CT-derived measurements (21–24), as detailed in Supplemental Table S1.

### Cardiovascular phenotypes and outcome classifications

Cardiovascular findings were grouped into phenotype domains based on established cardiovascular manifestations of thoracic radiation and cancer therapy (4–7,14). The comprehensive framework included pericardial phenotypes (4,5,10), myocardial phenotypes (4,8,13), coronary phenotypes (16,25–27), valvular phenotypes (6,11,15,28), great-vessel phenotypes (6,29,30), pulmonary vascular/right-heart phenotypes (7,14,30,31), and electrical phenotypes (5–7). Variables from different modalities representing the same pathological process were consolidated into a single phenotype to avoid duplication, e.g., pericardial effusion from CT or echocardiography and left ventricular ejection fraction (LVEF) from echocardiography or MPI. Domain-specific phenotypes, source modalities, and definitions are summarized in Supplemental Table S1. Variables considered but not included in the primary longitudinal analysis are summarized in Supplemental Table S2.

The primary outcome was temporal evolution of cardiovascular imaging and ECG phenotypes following definitive CRT. Secondary outcomes were clinical outcomes, including ischemic CVAEs (myocardial infarction and new coronary artery disease), nonischemic CVAEs (heart failure, clinically significant pericardial diseases, and clinically significant arrhythmia), and all-cause mortality.

Clinical cardiovascular adverse events (CVAEs) were classified as ischemic or nonischemic. Clinical CVAEs were coded by patient and follow-up interval, with at least one event in a CVAE category counted once for that category within an interval. Patients could contribute outcomes in multiple intervals or in both CVAE categories. Events were assigned to the follow-up interval in which they occurred.

### Statistical analysis

Baseline characteristics were summarized descriptively. Prevalence of new or progressive abnormalities at each follow-up interval was calculated using variable-specific denominators (Supplemental Table S3) and visualized with a temporal heatmap.

Longitudinal classifications were dichotomized as stable/improved = 0 and new/worsened = 1 compared to the baseline and then analyzed using population-averaged generalized estimating equation (GEE) models with logit link and patient-level clustering. Time was modeled ordinally across five categories post RT: 0-6 months = 1, 6-12 months = 2, 1-2 years = 3, 2-5 years = 4, and >5 years = 5. Odds ratios represent average change in odds per successive follow-up category. Univariable models included time as the sole variable. Multivariable models adjusted for age, sex, total radiation dose, and number of radiation fractions. Covariates were selected based on clinical relevance, treatment exposure, and consistency with overlapping cohort analysis (7,14,20). Models used all available observations without imputation. Variables with no new or worsening abnormalities during follow-up or insufficient data were excluded from GEE analyses.

Exploratory associations between cardiovascular phenotypes and clinical outcomes were assessed using Pearson’s χ² or Fisher’s exact tests, as appropriate. Clinical CVAEs were coded as binary outcomes (0 = absent, 1 = present) for each patient within each follow-up interval. Patients experiencing multiple events within the same CVAE category during an interval were counted once for that category, whereas patients experiencing both ischemic and nonischemic CVAEs could be classified as positive for both outcomes. These interval-specific analyses were considered exploratory and hypothesis-generating.

Interobserver agreement was assessed in a 15-case subset independently reviewed by the diagnostic radiology resident and fellowship-trained cardiothoracic radiologist using standardized definitions. Cohen κ with 95% CIs and exact agreement were reported (32). For variables with no category variation, κ was considered not estimable.

All tests were two-sided, and a P value ≤ 0.05 was considered statistically significant. Analyses were performed using R version 4.4.1 and JMP Student Edition version 19.1.1.

## RESULTS

### Baseline cohort characteristics

Baseline demographic and clinical characteristics are summarized in Table 1. The study included 86 patients with a mean age of 66.0 ± 9.1 years, 40 men and 46 women, and 83 White patients (96.5%). Most patients were former (66.3%) or current (29.1%) smokers, with a mean smoking history of 47.8 ± 30.2 pack-years. Hypertension (66.3%) and coronary artery disease (34.9%) were the most common baseline cardiovascular comorbidities. Squamous cell carcinoma (45.3%) was the predominant histologic subtype, followed by adenocarcinoma (29.1%) and small cell carcinoma (19.8%). Most patients had stage III disease (66.3%), received systemic therapy (98.8%) and immunotherapy (76.7%), and underwent IMRT/VMAT (98.8%).

**Table 1.** Baseline demographic, clinical, oncologic, and treatment characteristics of the study cohort.

| Characteristic | Overall (N = 86) | Characteristic | Overall (N = 86) |
|---|---|---|---|
| **Demographics** |  | **T Stage** |  |
| Age, mean ± SD (years) | 66.0 ± 9.1 | T1 | 20 (23.3%) |
| **Sex** |  | T2 | 16 (18.6%) |
| Female | 46 (53.5%) | T3 | 19 (22.1%) |
| Male | 40 (46.5%) | T4 | 21 (24.4%) |
| **Race** |  | Tx | 4 (4.7%) |
| White | 83 (96.5%) | N/A | 6 (7.0%) |
| Black | 3 (3.5%) | **N Stage** |  |
| **Smoking History** |  | N0 | 9 (10.5%) |
| Never | 4 (4.7%) | N1 | 9 (10.5%) |
| Former | 57 (66.3%) | N2 | 33 (38.4%) |
| Current | 25 (29.1%) | N3 | 25 (29.1%) |
| Pack-years, mean ± SD (years) | 47.8 ± 30.2 | Nx | 4 (4.7%) |
| **Clinical Characteristics** |  | N/A | 6 (7.0%) |
| **Baseline Cardiovascular Comorbidities** |  | **M Stage** |  |
| Congestive heart failure | 7 (8.1%) | M0 | 60 (69.8%) |
| Coronary artery disease | 30 (34.9%) | M1a | 0 (0.0%) |
| Hypertension | 57 (66.3%) | M1b | 4 (4.7%) |
| Cerebrovascular accident | 4 (4.7%) | M1c | 2 (2.3%) |
| Peripheral arterial disease | 9 (10.5%) | Mx | 14 (16.3%) |
| Conduction disorder/arrhythmia | 16 (18.6%) | N/A | 6 (7.0%) |
| Total comorbidities, mean ± SD | 2.64 ± 1.85 | **ECOG Performance Status** |  |
| **Primary Tumor Location** |  | 0 | 19 (22.1%) |
| Right lung | 43 (50.0%) | 1 | 43 (50.0%) |
| Left lung | 42 (48.8%) | 2 | 15 (17.4%) |
| Mediastinum | 1 (1.2%) | 3 | 2 (2.3%) |
| **Histology** |  | Unknown | 7 (8.1%) |
| Adenocarcinoma | 25 (29.1%) | **Non-Radiation Cancer Therapies** |  |
| Squamous cell carcinoma | 39 (45.3%) | Received systemic therapy | 85 (98.8%) |
| Small cell carcinoma | 17 (19.8%) | Received immunotherapy | 66 (76.7%) |
| Large cell carcinoma | 1 (1.2%) | Surgery | 12 (14.0%) |
| Poorly differentiated | 3 (3.5%) | **Radiation Characteristics** |  |
| Other | 1 (1.2%) | **RT Location** |  |
| **Clinical Stage** |  | Right lung | 41 (47.7%) |
| Stage I | 1 (1.2%) | Left lung | 35 (40.7%) |
| Stage II | 5 (5.8%) | Bilateral | 0 (0.0%) |
| Stage III | 57 (66.3%) | Mediastinum | 3 (3.5%) |
| Stage IV | 6 (7.0%) | Right lung + mediastinum | 5 (5.8%) |
| Limited stage SCLC | 17 (19.8%) | Left lung + mediastinum | 2 (2.3%) |
|  |  | **RT Technique** |  |
|  |  | 3DCRT | 1 (1.2%) |
|  |  | IMRT/VMAT | 85 (98.8%) |
|  |  | Prescription dose, Gy, median (range) | 60 (44–70) |
|  |  | Number of fractions, median (range) | 30 (22–35) |

*Note. Data are presented as n (%) or mean ± SD unless otherwise indicated. Abbreviations: 3DCRT = three-dimensional conformal radiation therapy; ECOG = Eastern Cooperative Oncology Group; IMRT = intensity-modulated radiation therapy; RT = radiation therapy; SCLC = small cell lung cancer; SD = standard deviation; VMAT = volumetric modulated arc therapy.*

### Baseline cardiovascular imaging and ECG findings

Baseline imaging and ECG findings are summarized in Table 2. Calcific phenotypes predominated: thoracic aortic calcification in 76/86 (88%), coronary artery calcification in 62/86 (72%), and valvular calcification in 43/86 (50%). Myocardial abnormalities included diastolic dysfunction in 18/45 (40%) patients, cardiomegaly in 18/84 (21%), global hypokinesis in 5/45 (11%), and left ventricular dilation in 5/48 (10%), with a mean LVEF of 59% ± 11.3%. Valvular dysfunction was present in 27/39 (69%) patients, including mild valvular dysfunction in 21/39 (54%) patients and moderate or severe dysfunction in 6/39 (15%). Pericardial effusion was present in 8/85 (9%) patients, without pericardial thickening. Main pulmonary artery enlargement was observed in 27/84 (32%) patients, while pulmonary hypertension, right ventricular dilation, conduction abnormalities, and arrhythmias were less common.

**Table 2.** Baseline multimodality cardiovascular imaging and electrocardiographic findings before definitive chemoradiation therapy.

| Phenotype Domain | Baseline Characteristic | Modality | Baseline Status |
|---|---|---|---|
| **Pericardial disease** | Pericardial effusion | CT/Echo | 8/85 (9%) |
|  | Pericardial thickening | CT | 0/82 (0%) |
| **Myocardial dysfunction & remodeling** | LVEF (average ± SD) | Echo/MPI | 59 ± 11.3% (n = 50) |
|  | Regional wall motion abnormality | Echo | 1/44 (2%) |
|  | Global hypokinesis | Echo | 5/45 (11%) |
|  | Diastolic dysfunction | Echo | 18/45 (40%) |
|  | LV dilation | Echo | 5/48 (10%) |
|  | Cardiomegaly | CT | 18/84 (21%) |
| **Coronary artery disease** | Coronary artery calcification | CT | - None: 24/86 (28%)<br>- Mild: 20/86 (23%)<br>- Moderate: 4/86 (5%)<br>- Severe: 38/86 (44%) |
|  | Perfusion abnormality | MPI | - Normal: 11/18 (61%)<br>- Reversible defect: 5/18 (28%)<br>- Fixed defect: 2/18 (11%) |
|  | Significant coronary stenosis (≥50%)** | Coronary angiography | 7/12 (58%) |
| **Valvular heart disease** | Valvular calcification | CT | 43/86 (50%) |
|  | Valvular dysfunction*** | Echo | - None/Trace: 12/39 (31%)<br>- Mild: 21/39 (54%)<br>- Moderate/Severe: 6/39 (15%) |
| **Great vessel disease** | Thoracic aortic calcification | CT | - None/Trace: 10/86 (12%)<br>- Mild: 47/86 (55%)<br>- Moderate: 12/86 (14%)<br>- Severe: 17/86 (20%) |
| **Pulmonary vascular / right heart disease** | Main pulmonary artery enlargement | CT | 27/84 (32%) |
|  | Pulmonary hypertension | Echo | - None: 33/39 (85%)<br>- Mild: 2/39 (5%)<br>- Moderate: 3/39 (8%)<br>- Severe: 1/39 (3%) |
|  | Right heart strain | CT | 9/61 (15%) |
|  | RV dilation | Echo | 4/38 (11%) |
| **Electrical abnormalities** | Conduction abnormality | ECG | 18/78 (23%) |
|  | Arrhythmia | ECG | 8/77 (10%) |

*Note. Cardiovascular findings across modalities were grouped into predefined cardiovascular phenotype domains. Data are presented as mean ± SD for continuous variables and n/N (%) for categorical variables unless otherwise indicated. Denominators represent the number of evaluable patients for each variable and differ according to source modality availability.*
*\*\*Coronary angiography was performed only when clinically indicated; therefore, the prevalence of significant coronary artery stenosis reflects the subgroup undergoing angiography rather than the entire cohort.*
*\*\*\*Valvular dysfunction reflects the highest-grade stenosis or regurgitation involving any cardiac valve for each patient.*
***Abbreviations:*** *CT = computed tomography; ECG = electrocardiography; Echo = echocardiography; LV = left ventricular; LVEF = left ventricular ejection fraction; MPI = myocardial perfusion imaging; RV = right ventricular; SD = standard deviation.*

### Temporal cardiovascular phenotypes

CT remained the most consistently available modality across follow-up intervals (Supplemental Table S3). Descriptive longitudinal assessment identified overlapping cardiovascular phenotype patterns across follow-up intervals (Figure 2): myocardial abnormalities, pericardial effusion, valvular dysfunction, and electrical abnormalities were observed within the first year; pulmonary vascular/right-heart abnormalities and calcification progression were more frequently documented during later follow-up. Representative longitudinal imaging examples are illustrated in Figure 3.

**Figure 2.**
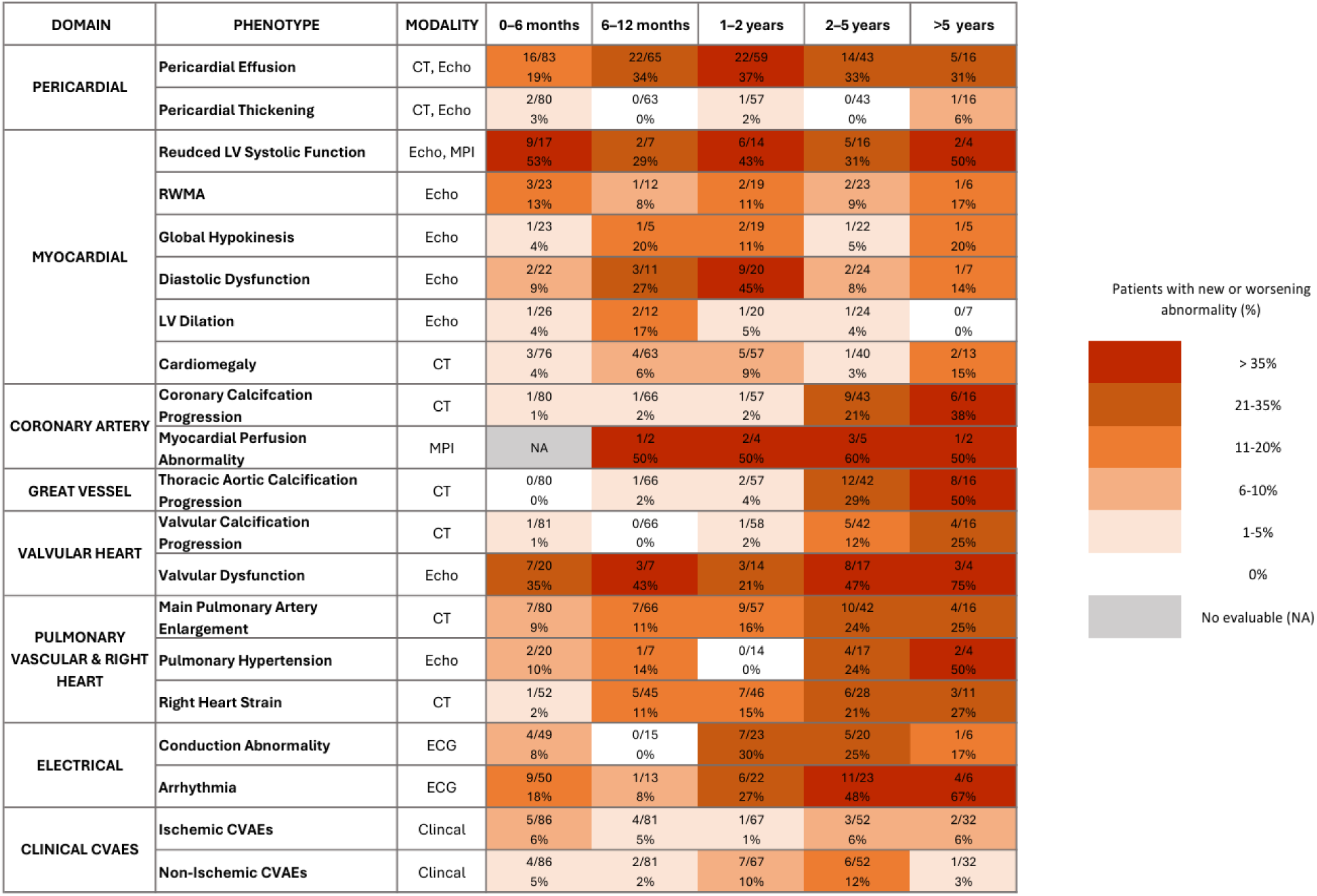
Temporal evolution of multimodality cardiovascular phenotypes following definitive chemoradiation therapy for lung cancer. Heatmap shows the number of evaluable patients with new or worsening cardiovascular imaging or electrocardiographic abnormalities (n/N), with the corresponding percentage displayed beneath, across five follow-up intervals after chemoradiation therapy. Percentages were calculated using variable-specific denominators according to source modality availability. Darker shading indicates higher prevalence within each interval. Clinical CVAEs are shown as outcome categories rather than imaging phenotypes; values represent patients with at least one event in the specified CVAE category within each interval. **Abbreviations:** CT = computed tomography; CVAE = cardiovascular adverse event; ECG = electrocardiography; Echo = echocardiography; LV = left ventricular; MPI = myocardial perfusion imaging; PA = pulmonary artery; RWMA = regional wall motion abnormality; RV = right ventricular

**Figure 3.**
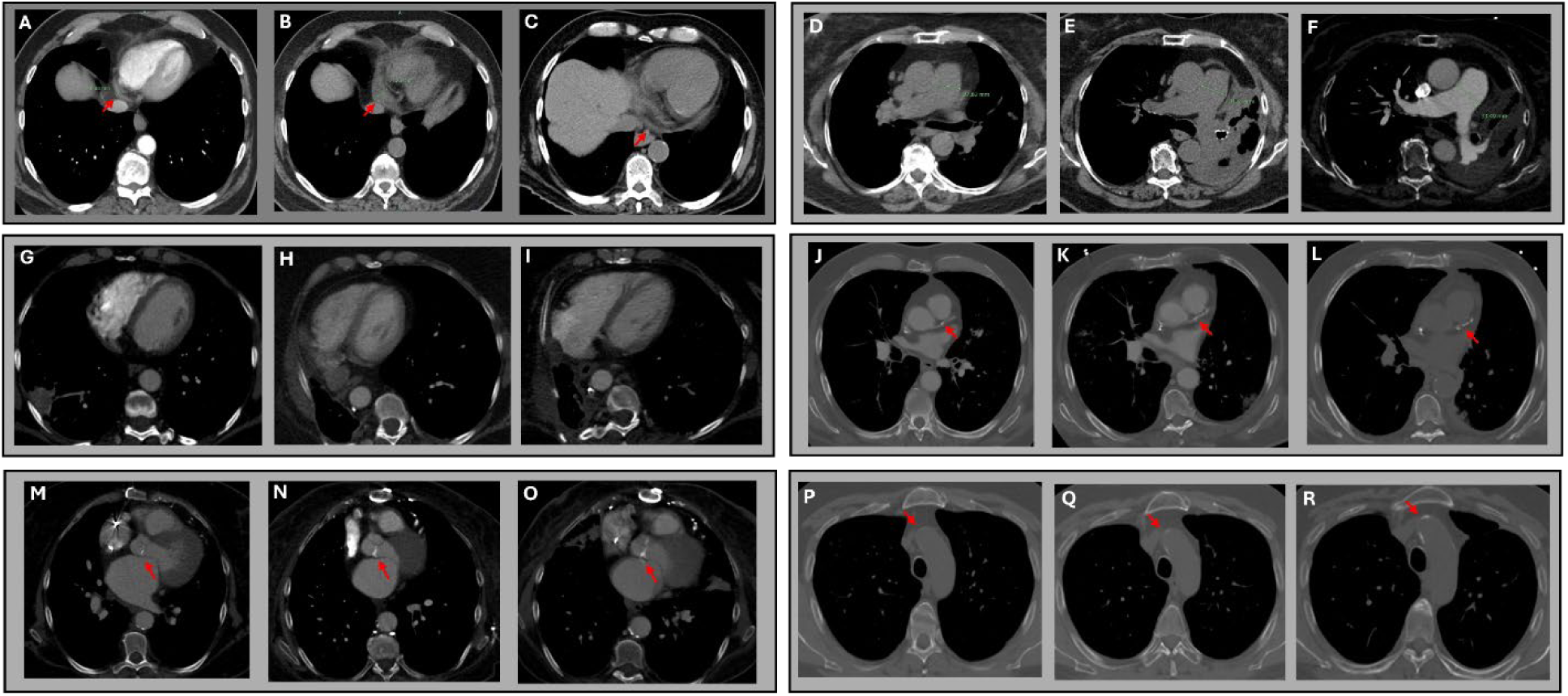
Representative longitudinal CT examples after definitive chemoradiotherapy; intervals are relative to radiotherapy start. (A–C) Pericardial effusion in a man with left-sided squamous cell carcinoma, measuring 6 mm at baseline (A; contrast-enhanced), 21 mm at 0–6 months (B; noncontrast), and trace beyond 5 years (C; noncontrast). (D–F) Main pulmonary artery enlargement in a woman with left-sided lung carcinoma, measuring 27–28 mm at baseline (D; noncontrast), 31–32 mm at 2–5 years (E; noncontrast), and 33–34 mm beyond 5 years (F; contrast-enhanced). (G–I) Increased RV/LV di-ameter ratio on contrast-enhanced CT in a woman with right-sided squamous cell carcinoma, from <1.0 at baseline (G) to 1.10 (RV, 45 mm; LV, 41 mm) at 1–2 years (H) and 1.05 (RV, 40 mm; LV, 38 mm) at 2–5 years (I). (J–L) Progressive left anterior descending coronary artery calcification in a man with left-sided squamous cell carcinoma at baseline (J) and 2–5 years (K) (contrast-enhanced) and beyond 5 years (L; noncontrast). (M–O) Progressive aortic valve calcification in the patient shown in A–C at baseline (M), 1–2 years (N), and 2–5 years (O) (contrast-enhanced). (P–R) Progressive thoracic aortic calcification in a man with left-sided adenocarcinoma at baseline (P), 1–2 years (Q), and 2–5 years (R) (contrast-enhanced). Arrows indicate the abnormality.

Longitudinal GEE analyses observed significant monotonic temporal patterns across several phenotypes (Table 3). In univariable models, increasing follow-up category was associated with higher odds of LV dilation (OR, 3.29; 95% CI: 1.06–10.21; P = 0.039), progressive coronary calcification (OR, 3.46; 95% CI: 1.82-6.57; P < 0.001), thoracic aortic calcification (OR, 5.00; 95% CI: 2.92-8.56; P < 0.001), valvular calcification (OR, 3.17; 95% CI: 1.58-6.35; P = 0.001), pulmonary artery enlargement (OR, 1.46; 95% CI: 1.09-1.96; P = 0.011), right-heart strain (OR, 1.76; 95% CI: 1.26-2.45; P = 0.001), conduction abnormalities (OR, 1.47; 95% CI: 1.07-2.01; P = 0.017), and arrhythmias (OR, 1.68; 95% CI: 1.25-2.25; P < 0.001). These significant univariable phenotypes were carried forward into multivariable models.

**Table 3.**
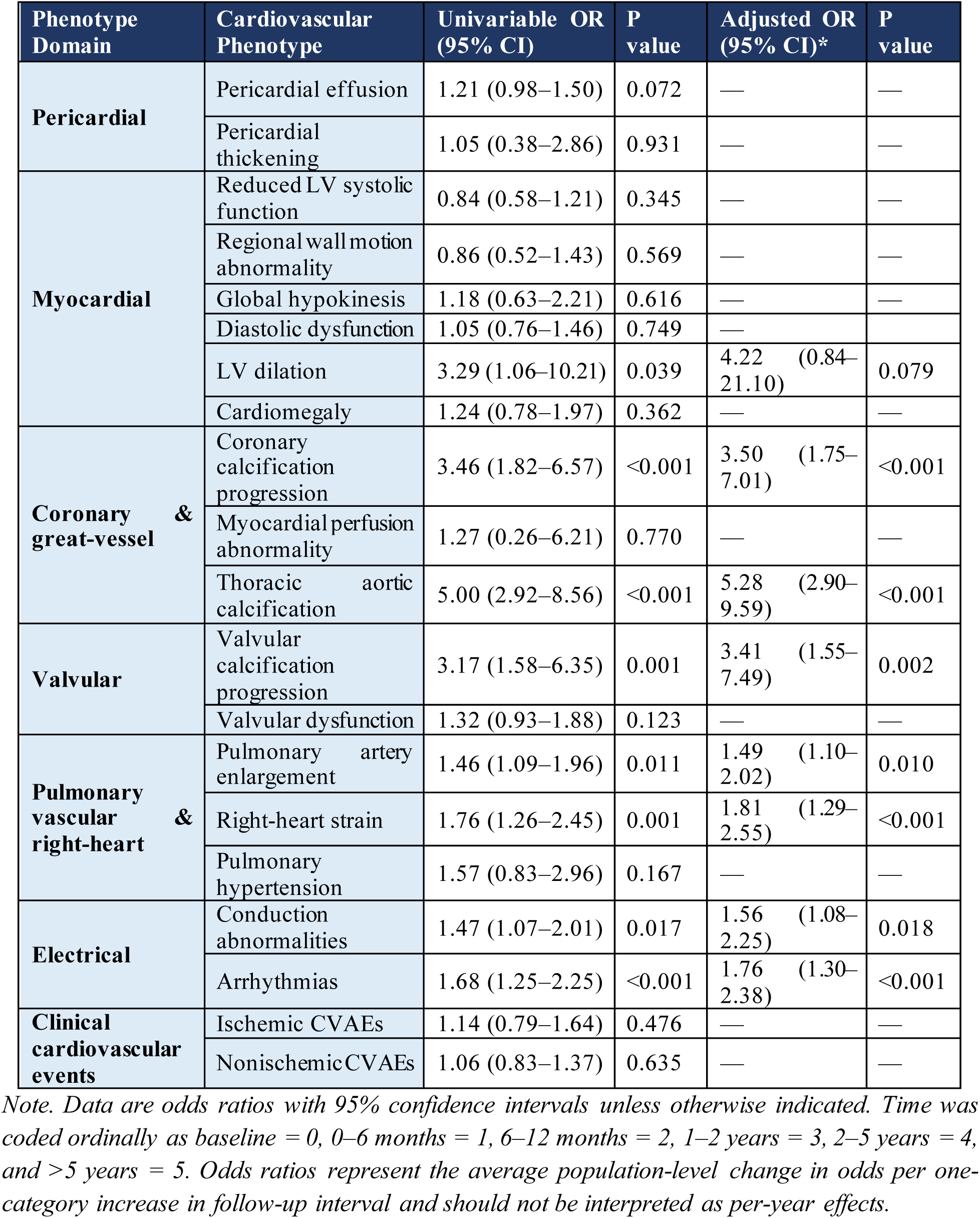
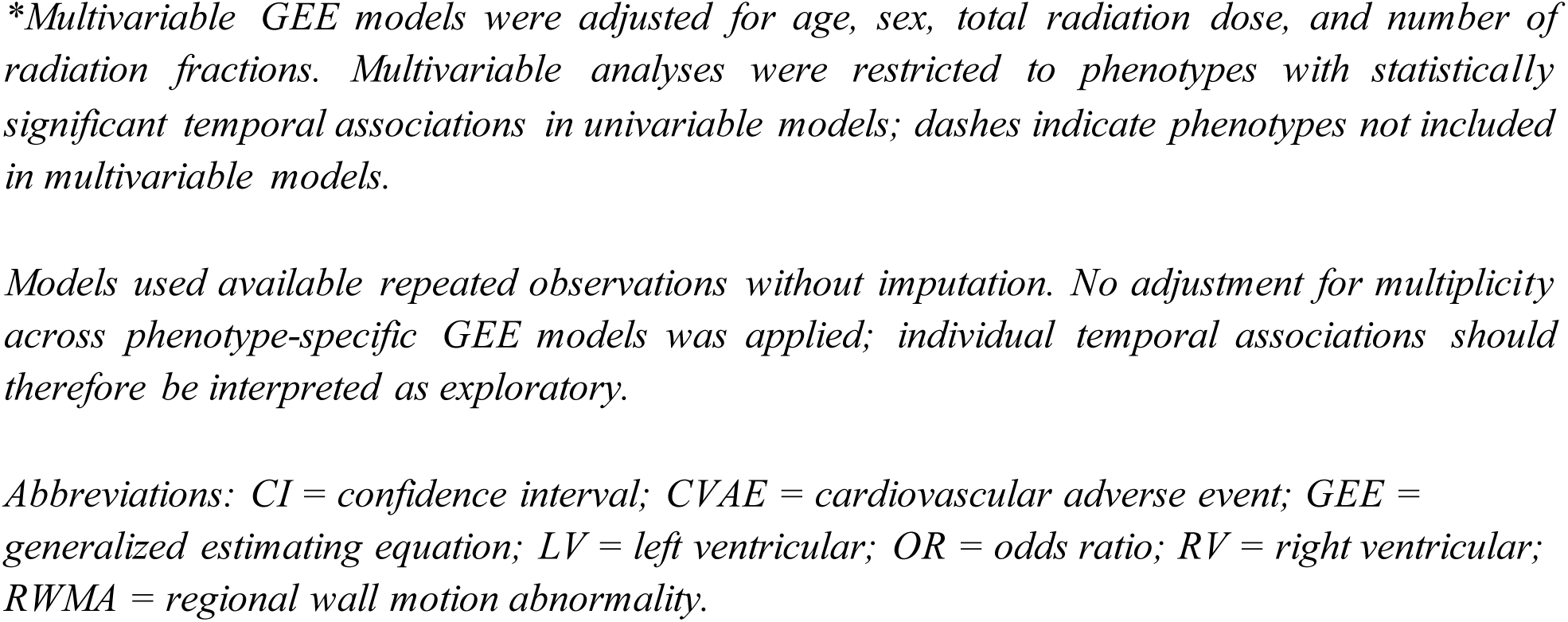
Univariable and multivariable GEE analyses of temporal trends in cardiovascular phenotypes and clinical CVAEs.

In multivariable models, increasing odds across successive follow-up categories remained significant for thoracic aortic calcification (OR, 5.28; 95% CI: 2.90-9.59; P < 0.001), coronary calcification (OR, 3.50; 95% CI: 1.75-7.01; P < 0.001), valvular calcification (OR, 3.41; 95% CI: 1.55-7.49; P = 0.002), right-heart strain (OR, 1.81; 95% CI: 1.29-2.55; P < 0.001), arrhythmias (OR, 1.76; 95% CI: 1.30-2.38; P < 0.001), conduction abnormalities (OR, 1.56; 95% CI: 1.08-2.25; P = 0.018), and pulmonary artery enlargement (OR, 1.49; 95% CI: 1.10-2.02; P = 0.010). LV dilation was significant in univariable analysis but did not remain significant after multivariable adjustment. Reduced LV systolic function, pericardial effusion, valvular dysfunction, and clinical CVAEs remained prevalent but did not demonstrate significant monotonic temporal trends. Observed means of cardiovascular phenotypes with significant monotonic temporal trends are shown in Supplemental Figure S1.

### Clinical cardiovascular events

Clinical CVAEs were summarized as binary patient-interval outcomes in Figure 2. The observed proportions varied across follow-up intervals (ischemic CVAEs, 1%-6%; nonischemic CVAEs, 2%-12%), with event ascertainment based on available clinical follow-up.

### Phenotype-outcome associations

Exploratory phenotype-outcome association analyses are summarized as a complete P-value matrix in Supplemental Table S4. Most imaging phenotypes were not significantly associated with clinical outcomes.

Significant associations were most evident between electrical abnormalities and nonischemic CVAEs during early and intermediate follow-up. At 0–6 months, arrhythmia and conduction abnormality were associated with nonischemic CVAEs (P < 0.001 and P = 0.003, respectively). At 1-2 years, arrhythmia and conduction abnormality remained associated with nonischemic CVAEs (P = 0.007 and P < 0.001, respectively).

During later follow-up, RWMA showed the broadest clinical associations. At 2-5 years, RWMA was associated with ischemic CVAEs (P = 0.005), nonischemic CVAEs (P = 0.050), and all-cause mortality (P = 0.013). Additional significant associations included LV dilation with ischemic CVAEs at 6-12 months (P = 0.045), right-heart strain with nonischemic CVAEs at 6-12 months (P = 0.002), thoracic aortic calcification with ischemic CVAEs at 1-2 years (P < 0.001), and valvular calcification with nonischemic CVAEs at 2-5 years (P = 0.048). No significant associations were identified beyond 5 years.

### Interobserver agreement

Interobserver agreement for imaging phenotype classification is summarized in Supplemental Table S5. Across the 15-case reliability subset, the exact agreement ranged from 86.7% to 100%. Cohen κ demonstrated substantial-to-perfect agreement for pericardial effusion and calcific phenotypes (κ = 0.722-1.000) and moderate agreement for pulmonary artery enlargement (κ = 0.444). For cardiomegaly, myocardial thinning, and pericardial thickening, κ was not estimable because both readers assigned all cases to the same category. For findings with only a single discordant positive rating, κ was limited despite high exact agreement.

## DISCUSSION

### Principal findings

Using a longitudinal multimodality cardiovascular phenotype approach, our study demonstrated that cardiovascular abnormalities following definitive CRT for lung cancer exhibit heterogeneous longitudinal patterns across cardiovascular imaging and ECG phenotype domains. Early survivorship was characterized predominantly by myocardial, pericardial, and electrical phenotypes, whereas pulmonary vascular/right-heart abnormalities and calcific progression became prominent during intermediate-to-late follow-up. Multivariable GEE analyses demonstrated significant monotonic temporal increases in calcific, pulmonary vascular/right-heart, and electrical phenotypes, whereas myocardial and pericardial phenotypes remained persistent without significant monotonic progression. These findings extend beyond prior studies, which largely focused on individual cardiovascular abnormalities, single imaging modalities, or isolated clinical endpoints. The multimodality approach used in our study provided complementary and, when appropriate, confirmatory characterization of cardiovascular findings; for example, echocardiography could help distinguish or confirm pericardial effusion when CT findings were equivocal for pericardial effusion versus thickening. A comprehensive multimodality framework characterizing the temporal evolution of cardiovascular phenotypes after thoracic CRT is proposed in Figure 4.

**Figure 4.**
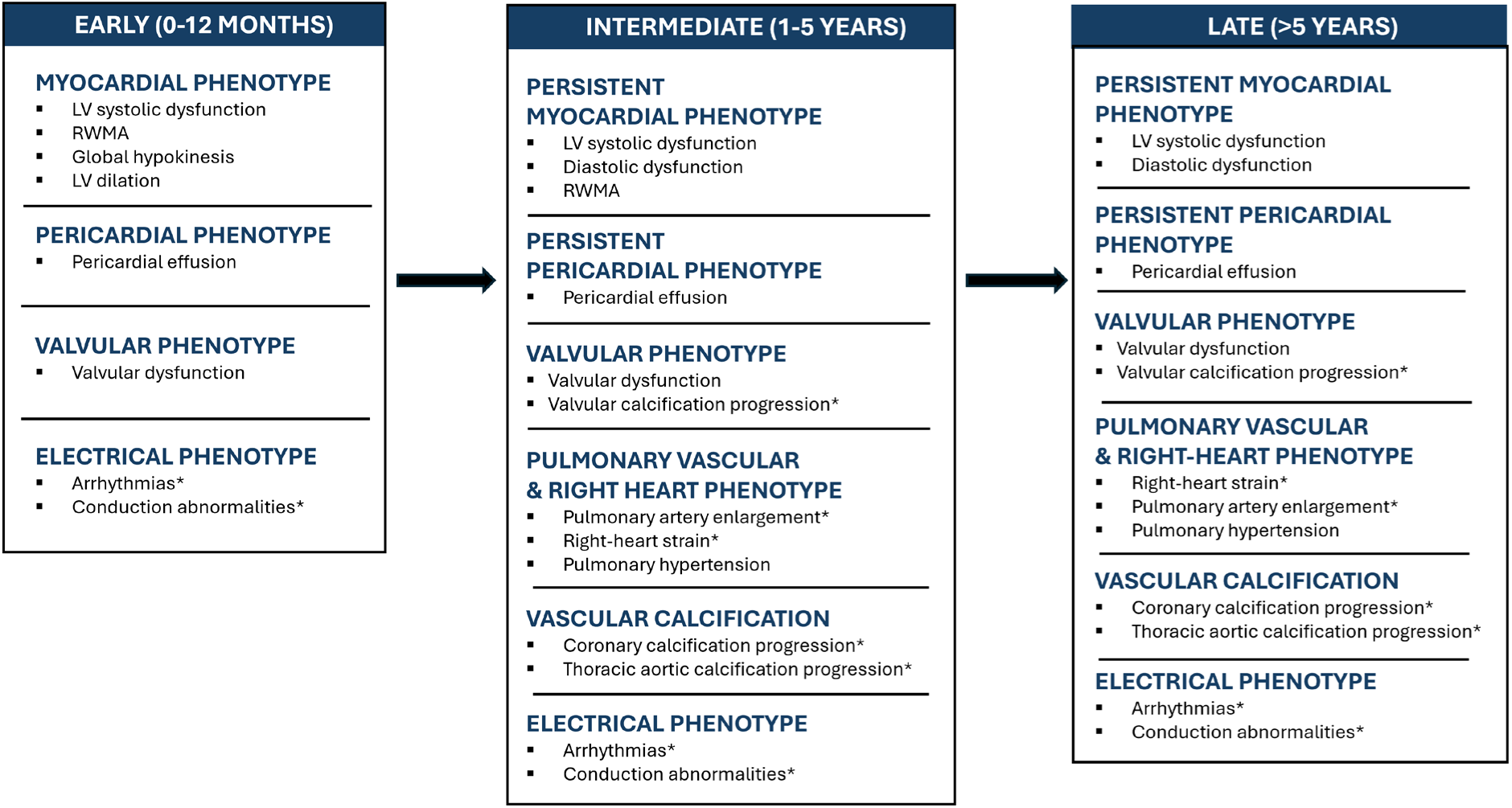
Proposed framework for temporal cardiovascular phenotype evolution following definitive chemoradiation therapy for lung cancer. Conceptual schematic summarizes predominant cardiovascular phenotype domains during early (0-12 months), intermediate (1-5 years), and late (>5 years) survivorship based on longitudinal multimodality imaging and electrocardiographic findings. Repetition across phases indicates persistence at the cohort level and does not imply continuous progression within individual patients. “Persistent” indicates continued presence across multiple intervals without necessarily demonstrating a monotonic increase. *Phenotype demonstrated significantly increasing odds across successive follow-up categories in multivariable generalized estimating equation (GEE) models. Abbreviations: CRT = chemoradiation therapy; GEE = generalized estimating equation; LV = left ventricular; RWMA = regional wall motion abnormality.

Because lung cancer patients frequently have advanced age, heavy tobacco exposure, chronic obstructive pulmonary disease, pre-existing cardiovascular comorbidity, and competing cardiovascular risks, these imaging and ECG phenotypes should be interpreted as multifactorial rather than solely treatment-related (7,31).

### Temporal patterns across phenotype domains

Several phenotype domain-specific patterns are particularly relevant to imaging interpretation. Pericardial phenotype domain was an early and persistent manifestation, driven predominantly by pericardial effusion, consistent with prior studies reporting pericardial effusion as an early manifestation after definitive CRT (5,10,28). Myocardial phenotypes predominated early and remained evident throughout follow-up, consistent with prior multimodality studies (11–13), but showed sustained or fluctuating rather than monotonic accumulation. The limited availability of cardiac MRI restricted tissue-level characterization of fibrosis, edema, and inflammation (9). Calcific phenotypes were readily assessable on routine chest CT and showed the most apparent delayed progression of coronary, thoracic aortic, and valvular calcification. This pattern is consistent with prior reports of delayed vascular effects of thoracic irradiation (16,26) and the recognized pathophysiology of accelerated atherosclerosis (5,29). Valvular dysfunction by echocardiography remained prevalent throughout survivorship but did not parallel the monotonic temporal trend of valvular calcification by CT, underscoring the complementary roles of CT and echocardiography. Pulmonary artery enlargement and right-heart strain also progressively increased over time, consistent with previous studies linking thoracic radiotherapy with pulmonary vascular remodeling, pulmonary hypertension, and right ventricular loading conditions (29,31). Electrical phenotypes showed progressive temporal evolution and demonstrated the most consistent interval-specific associations with CVAEs, supporting ECG as a potential complementary longitudinal surveillance tool.

### Phenotype-outcome associations and clinical implications

The exploratory phenotype-outcome association analyses are hypothesis-generating rather than confirmatory because they are unadjusted, interval-specific, uncorrected for multiple comparisons, and limited in some cases by sparse events and zero-cell counts. However, they provide clinical context that may help prioritize cardiovascular imaging phenotypes for future prognostic validation. Given their consistent association with nonischemic CVAEs, electrical phenotypes may provide a readily available complement to imaging surveillance. RWMA may represent a late-predominant myocardial phenotype with potential clinical relevance to ischemic events and all-cause mortality. However, all-cause mortality in lung cancer requires caution because it is influenced by tumor biology, stage, disease progression, baseline comorbidity, and competing non cardiovascular risks.

Imaging and ECG abnormalities were substantially more frequent than clinically recognized CVAEs, indicating that routine multimodality oncologic and cardiovascular imaging and ECG may identify cardiovascular risks before or without overt clinical events (7,28). These findings are not diagnostic of treatment-related injury but may identify patients who warrant closer multidisciplinary surveillance. Standardized cardiovascular phenotype definitions may also inform imaging endpoints for future prediction studies (33).

To our knowledge, this study is among the first to integrate longitudinal multimodality imaging and ECG findings across cardiovascular phenotype domains after definitive CRT for lung cancer, with follow-up extending beyond five years. Standardized follow-up intervals, repeated-measures GEE analysis, and interobserver agreement assessment enhanced analytic transparency, while inclusion of an underrepresented Appalachian cohort with substantial cardiovascular risks expands the limited longitudinal cardio-oncology literature in this population.

### Limitations and future directions

Limitations include retrospective single-center design, clinically acquired rather than protocolized imaging, variable availability of imaging modalities over time, potential informative missingness, and residual confounding. In particular, cardiac MRI was insufficient for analysis, precluding comprehensive myocardial tissue characterization. The proposed framework reflects population-level temporal patterns rather than individual biological progression and does not establish causality.

Future studies should prospectively validate the proposed temporal cardiovascular phenotype framework in larger multicenter cohorts using standardized multimodality imaging protocols, ECG, biomarkers, cardiac MRI, radiation dosimetry, clinical risk factors, and prediction models to further refine individualized cardio-oncology surveillance and improve early detection of clinically significant cardiovascular injury.

## Conclusion

Cardiovascular abnormalities after definitive CRT for lung cancer exhibited heterogeneous, overlapping longitudinal phenotype patterns. Longitudinal multimodality imaging and ECG provide complementary characterization of these changes and a framework for prospective validation of phenotype-based cardiovascular surveillance.

## Data Availability

All data produced in the present study are available upon reasonable request to the authors

## Ethics and Consents

The study was approved by West Virginia University Institutional Board Review (WVU Protocol #: 2308839854, Approved on 01/09/2024) which waived the need for consent forms due to retrospective nature of the study.

## Data sharing statement

De-identified clinical and imaging materials will be available upon request from the corresponding author according to the institutional restricted data sharing and IRB requirements.

## Author contributions

Troy Nguyen – Conceptualization, Data curation, Formal analysis, Investigation, Methodology, Project administration, Validation, Visualization, Writing – original draft, Writing – review & editing

Vivian Salama – Conceptualization, Data curation, Formal analysis, Funding acquisition, Investigation, Methodology, Project administration, Supervision, Validation, Writing – review & editing

Joseph A. Schmidlen – Data curation, Writing – original draft, Writing – review & editing Peyton M. Lilly – Data curation, Writing – original draft

Sijin Wen – Formal analysis, Methodology, Validation

Nathaniel Dunham – Writing – original draft, Writing – review & editing

R Alfredo Siochi – Data curation, Writing – review & editing

Bavly Naklah – Writing – original draft

David A. Clump, MD – Supervision, Resources

Phillip M. Pifer – Supervision, Resources

Mina F. Hanna – Conceptualization, Investigation, Methodology, Supervision, Validation, Writing – review and editing

## Acknowledgments

We acknowledge the West Virginia Clinical and Translational Science Institute and Tyler McGaughey, PhD, for assistance with imaging file extraction, anonymization, and secure institutional repository deposition.

## Abbreviations

CRT: Chemoradiation therapy
CT: Computed tomography
CVAE: Cardiovascular adverse event
ECG: Electrocardiography
GEE: Generalized estimating equation
LV: Left ventricle
LVEF: Left ventricular ejection fraction
MRI: Magnetic resonance imaging
NSCLC: Non-small cell lung cancers
RWMA: Regional wall motion abnormality

