## Supplemental Files for "Longitudinal Multimodality Imaging of Cardiovascular Abnormalities Following Definitive Chemoradiation for Lung Cancer: A Retrospective Cohort Study"

| Phenotype Domains | Cardiovascular Phenotypes | Source Modality | Definition/Measurement | Variable Format | Role in Primary Analysis |
|---|---|---|---|---|---|
| Pericardial | **Pericardial effusion** | CT, Echo | New or increased pericardial fluid on CT or echocardiography compared with baseline. If CT and echocardiography were discordant, the more severe abnormality was recorded. | Binary (Present/Absent) | Primary phenotype |
| | **Clinically significant pericardial effusion** | Clinical records | Pericardial effusion documented in the clinical record as a cardiovascular adverse event requiring clinical evaluation, management, or attributed to the patient's symptoms, regardless of whether formal diagnostic criteria for pericarditis were met. | Binary (Present/Absent) | No (Clinical CVAE) |
| | **Pericardial thickening** | CT, Echo | Pericardial thickness >2 mm or visually abnormal thickening on CT (21). Echocardiographic findings were extracted from final reports. | Binary (Present/Absent) | Primary phenotype |
| | **Pericarditis** | Clinical records | Clinically diagnosed pericarditis based on treating physician documentation. | Binary (Yes/No) | No (Clinical CVAE) |
| Myocardial | **LV systolic dysfunction** | Echo, MPI | Reduced resting LVEF compared with baseline. Echocardiographic LVEF was preferentially used when available; resting MPI LVEF used if Echo unavailable. Stress LVEF was not used. | Continuous (%) & binary (reduced vs preserved) | Primary phenotype |
| | **Global hypokinesis** | Echo | Diffuse reduction in LV wall motion reported on echocardiography. | Binary (Present/Absent) | Primary phenotype |
| | **Regional wall motion abnormality** | Echo | New or worsening regional wall motion abnormality. | Binary (Present/Absent) | Primary phenotype |
| | **Diastolic dysfunction** | Echo | New or worsening diastolic dysfunction of any grade. | Binary (Present/Absent) | Primary phenotype |
| | **LV dilation** | Echo | Left ventricular enlargement reported on echocardiography. | Binary (Present/Absent) | Primary phenotype |
| | **Cardiomegaly** | CT | Cardiothoracic ratio >0.5 or radiologist interpretation of cardiomegaly (22). | Binary (Present/Absent) | Primary phenotype |
| | **Myocardial thinning** | CT | Focal thinning of irradiated myocardium identified on nongated CT. | Binary (Present/Absent) | Excluded |
| | **Abnormal myocardial attenuation** | CT | Focal hypoattenuation of myocardium on routine CT not attributable to artifact. | Binary (Present/Absent) | Excluded |
| | **Global longitudinal strain (GLS)** | Echo | Reported GLS value when available. | Continuous (%) | Excluded |
| Coronary artery | **Coronary artery calcification** | CT | Baseline severity graded visually. | Ordinal (None/Mild/Moderate/Severe) | Baseline only |

| | | | | | |
|---|---|---|---|---|---|
| | **Coronary artery calcification progression** | CT | New or worsened coronary calcification compared with baseline. | Binary (Present/Absent) | Primary phenotype |
| | **Perfusion abnormality** | MPI | Reversible and/or fixed perfusion defect. Reversible and irreversible defects were combined into a single imaging phenotype. | Binary (Present/Absent) | Primary phenotype |
| | **Significant coronary stenosis** | Coronary angiography | ≥50% luminal stenosis in a major coronary artery. | Continuous (%) / ≥50% | Excluded |
| | **PCI/CABG** | Coronary angiography | Coronary revascularization performed after clinical diagnosis. | Binary (Yes/No) | Excluded |
| **Great vessel** | **Baseline thoracic aortic calcification** | CT | Baseline thoracic aortic calcification severity graded visually. | Ordinal (None/Mild/Moderate/Severe) | Baseline only |
| | **Thoracic aortic calcification progression** | CT | New or worsened thoracic aortic calcification. | Binary (Yes/No) | Primary phenotype |
| **Valvular** | **Baseline valvular calcification** | CT | Baseline valvular calcification severity graded visually. | Ordinal (None/Mild/Moderate/Severe) | Baseline only |
| | **Valvular calcification progression** | CT | New or worsened valvular calcification compared with baseline. | Binary (Yes/No) | Primary phenotype |
| | **Valvular dysfunction** | Echo | Highest severity among all valves (MR, AR, AS, MS, TR, PR). | Ordinal (None/Mild/Moderate/Severe) | Primary phenotype |
| **Pulmonary vascular & right-heart** | **Main pulmonary artery enlargement** | CT | Main PA diameter >29 mm in men, >27 mm in women, or interval enlargement (23). | Binary (Present/Absent) | Primary phenotype |
| | **Pulmonary hypertension** | Echo | Estimated RVSP/PA systolic pressure or interpreting cardiologist diagnosis. | Ordinal (None/Mild/Moderate/Severe) | Primary phenotype |
| | **Right heart strain** | CT | RV/LV ratio ≥ 1.0 (24) | Binary (Present/Absent) | Primary phenotype |
| | **RV dilation** | Echo | Right ventricular enlargement on echocardiography. | Binary (Present/Absent) | Primary phenotype |
| **Electrical** | **Conduction abnormality** | ECG | Bundle branch block, fascicular block, AV block, or other conduction delay. | Binary (Present/Absent) | Primary phenotype |
| | **Arrhythmia** | ECG | New atrial fibrillation/flutter, SVT, ventricular arrhythmia, or frequent ectopy. | Binary (Present/Absent) | Primary phenotype |

| | | | | | |
|---|---|---|---|---|---|
| **Clinical adverse events** | **Ischemic CVAE** | Clinical records | STEMI, NSTEMI, unstable angina, or symptomatic obstructive CAD. | Binary (Yes/No) | Clinical outcome |
| | **Non-ischemic CVAE** | Clinical records | Heart failure, clinically diagnosed pericarditis, clinically significant pericardial effusion, clinically significant arrhythmia, and pulmonary hypertension. | Binary (Yes/No) | Clinical outcome |
| | **Mortality** | Clinical records | Death from any cause during follow-up. | Binary (Alive/Dead) | Clinical outcome |

**Supplemental Table S1. Cardiovascular phenotype variable dictionary and coding schema.**

Note. This table summarizes variables included in the cardiovascular phenotype framework, source modalities, operational definitions, coding schemes, and inclusion in the primary longitudinal heatmap analysis.

For baseline analyses, continuous measurements such as LVEF were summarized using the preferred modality, with echocardiography favored over resting MPI when both were available. Categorical baseline variables, including coronary, valvular, and thoracic aortic calcification, were reported using predefined severity grades.

For longitudinal analyses, variables were assessed relative to baseline and classified as new/worsened, unchanged, or improved. When multiple examinations were available within a follow-up interval, the most severe new or worsened abnormality was recorded.

When multiple modalities evaluated the same pathologic process, findings were consolidated into a single cardiovascular phenotype using the most severe abnormality within each follow-up interval. Examples include pericardial effusion on CT or echocardiography and LV systolic function on echocardiography or MPI.

Clinical CVAEs and all-cause mortality were analyzed separately from imaging and electrocardiographic phenotypes to distinguish clinically recognized outcomes from subclinical cardiovascular abnormalities.

All CT examinations were routine nongated chest CT examinations performed as part of oncologic surveillance unless otherwise specified. CT-derived measurements and visual grading criteria were adapted from published imaging literature where applicable (21-24). Progression variables were defined as new or visually increased abnormalities compared with baseline. Electrocardiographic findings were abstracted from official cardiology interpretations.

Abbreviations: AR = aortic regurgitation; AS = aortic stenosis; AV = atrioventricular; CABG = coronary artery bypass grafting; CAD = coronary artery disease; CT = computed tomography; CVAE = cardiovascular adverse event; ECG = electrocardiography; Echo = echocardiography; GLS = global longitudinal strain; LV = left ventricular; LVEF = left ventricular ejection fraction; MI = myocardial infarction; MPI = myocardial perfusion imaging; MR = mitral regurgitation; MS = mitral stenosis; PA = pulmonary artery; PCI = percutaneous coronary intervention; PR = pulmonic regurgitation; RV = right ventricular; RVSP = right ventricular systolic pressure; SVT = supraventricular tachycardia; TR = tricuspid regurgitation.

| Variable | Source Modality | Reason for Exclusion | Included in Other Analyses |
|---|---|---|---|
| **Global longitudinal strain (GLS)** | Echo | Insufficient availability for meaningful longitudinal analysis | No |
| **Myocardial thinning in irradiated segments** | CT | Subjective assessment without standardized measurement on routine nongated CT | No |
| **Abnormal myocardial attenuation** | CT | Nonspecific finding on routine nongated CT | No |
| **RV dilation** | Echo | No new or worsening cases during follow-up | Yes (Baseline phenotype analysis) |
| **Main vessel coronary stenosis (%)** | Coronary angiography | Insufficient availability for meaningful longitudinal analysis | Yes (Baseline phenotype analysis) |
| **Individual valve-specific abnormalities (MR, AR, TR, PR, AS)** | Echo | Consolidated into the composite Valvular Dysfunction phenotype using the highest severity across all cardiac valves | Yes (Valvular dysfunction phenotype) |
| **LVEF with pharmacologic stress** | MPI | Resting LVEF or echocardiographic LVEF was preferentially used to represent baseline ventricular systolic function | Yes (Reduced Ventricular Systolic Function) |
| **Segmental perfusion defect (reversible vs irreversible)** | MPI | Reversible and irreversible defects were combined into a single Perfusion Abnormality variable | Yes (Perfusion Abnormality) |
| **Pericarditis (clinical diagnosis)** | Clinical records | Low event frequency (3 events) and substantial overlap with imaging-detected pericardial abnormalities. Clinical events were captured within nonischemic CVAEs | No |

**Supplemental Table S2. Candidate variables evaluated but excluded from the final longitudinal phenotype analysis.**

Note. Variables were excluded from the primary longitudinal phenotype analysis because of insufficient longitudinal availability, lack of standardized assessment, redundancy with composite variables, limited reproducibility, sparse observations, or limited clinical applicability.

Abbreviations: AR = aortic regurgitation; AS = aortic stenosis; CT = computed tomography; Echo = echocardiography; GLS = global longitudinal strain; LV = left ventricular; LVEF = left ventricular ejection fraction; MR = mitral regurgitation; MPI = myocardial perfusion imaging; PR = pulmonic regurgitation; RV = right ventricular; TR = tricuspid regurgitation.

| Domain | Phenotype | Modality source | Baseline | 0–6 mo | 6–12 mo | 1–2 yr | 2–5 yr | >5 yr |
|---|---|---|---|---|---|---|---|---|
| Pericardial | Pericardial effusion | CT, Echo | 85 | 83 | 65 | 59 | 43 | 16 |
| | Pericardial thickening | CT, Echo | 82 | 80 | 63 | 57 | 43 | 16 |
| Myocardial dysfunction & remodeling | Reduced LV systolic function | Echo, MPI | 50 | 17 | 7 | 14 | 16 | 4 |
| | Regional wall motion abnormality | Echo | 44 | 23 | 12 | 19 | 23 | 6 |
| | Global hypokinesis | Echo | 45 | 23 | 5 | 19 | 22 | 5 |
| | Diastolic dysfunction | Echo | 45 | 22 | 11 | 20 | 24 | 7 |
| | LV dilation | Echo | 48 | 26 | 12 | 20 | 24 | 7 |
| | Cardiomegaly | CT | 84 | 76 | 63 | 57 | 40 | 13 |
| Coronary artery | Coronary calcification progression | CT | 86 | 80 | 66 | 57 | 43 | 16 |
| | Perfusion abnormality | MPI | 18 | 0 | 2 | 4 | 5 | 2 |
| Valvular | Valvular calcification progression | CT | 86 | 81 | 66 | 58 | 42 | 16 |
| | Valvular dysfunction | Echo | 39 | 20 | 7 | 14 | 17 | 4 |
| Great vessel | Thoracic aortic calcification progression | CT | 86 | 80 | 66 | 57 | 42 | 16 |
| Pulmonary vascular/right heart | Main pulmonary artery enlargement | CT | 84 | 80 | 66 | 57 | 42 | 16 |
| | Pulmonary hypertension | Echo | 39 | 20 | 7 | 14 | 17 | 4 |
| | Right heart strain | CT | 61 | 52 | 45 | 46 | 28 | 11 |
| Electrical | Conduction abnormality | ECG | 78 | 49 | 15 | 23 | 20 | 6 |
| | Arrhythmia | ECG | 77 | 50 | 13 | 22 | 23 | 6 |
| Clinical CVAEs | Ischemic CVAEs | Clinical | NA | 86 | 81 | 67 | 52 | 32 |
| | Non-ischemic CVAEs | Clinical | NA | 86 | 81 | 67 | 52 | 32 |

**Supplemental Table S3. Phenotype-specific numbers of evaluable patients based on source modality availability across longitudinal follow-up intervals.**

Note. Data are numbers of evaluable patients for each cardiovascular variable at baseline and within each follow-up interval after definitive chemoradiation therapy. Denominators represent patients with available imaging studies, electrocardiograms, or clinical follow-up data for each variable and were used to calculate the n/N values and percentages shown in Figure 2. Differences in denominators reflect variation in clinical utilization and availability of individual modalities during longitudinal follow-up.

Baseline values represent evaluable patients before initiation of definitive chemoradiation therapy. For imaging and ECG variables, denominators indicate patients with an evaluable study within each follow-up interval. For ischemic and nonischemic CVAEs, denominators represent patients alive and under clinical follow-up during each interval.

A value of 0 indicates that no patients had an evaluable study for that variable during the corresponding interval. NA indicates that the variable was not applicable to that time point.

Abbreviations: CT = computed tomography; CVAE = cardiovascular adverse event; Echo = echocardiography; ECG = electrocardiography; LV = left ventricular; MPI = myocardial perfusion imaging; NA = not applicable.

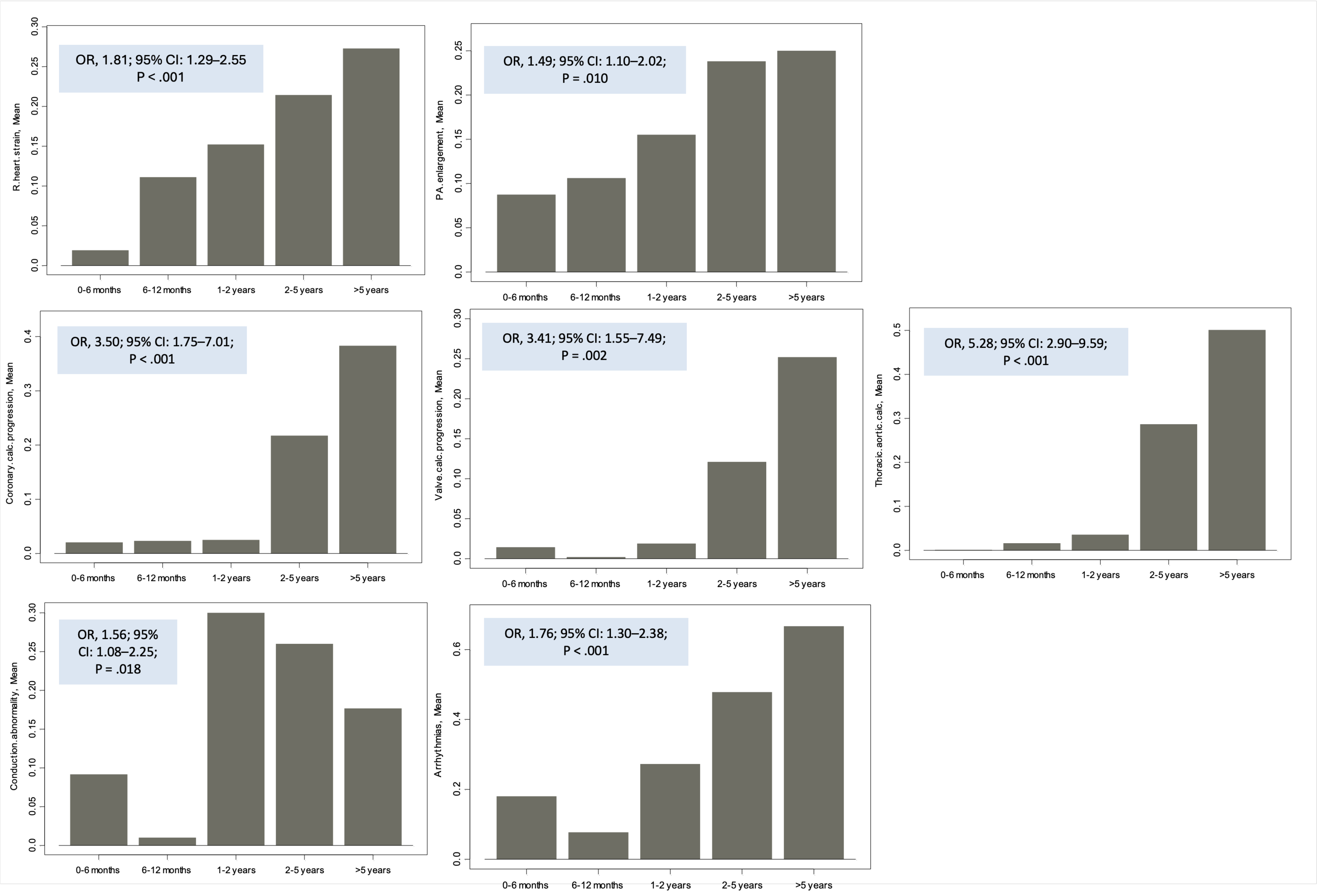
OR, 1.81; 95% CI: 1.29–2.55
P < .001
R.heart.strain, Mean
OR, 1.49; 95% CI: 1.10–2.02;
P = .010
P.A.enlargement, Mean
OR, 3.50; 95% CI: 1.75–7.01;
P < .001
Coronary.calc.progression, Mean
OR, 3.41; 95% CI: 1.55–7.49;
P = .002
Valve.calc.progression, Mean
OR, 5.28; 95% CI: 2.90–9.59;
P < .001
Thoracic.aortic.calc, Mean
OR, 1.56; 95%
CI: 1.08–2.25;
P = .018
Conduction.abnormality, Mean
OR, 1.76; 95% CI: 1.30–2.38;
P < .001
Arrhythmias, Mean
0-6 months
6-12 months
1-2 years
2-5 years
>5 years

**Supplemental Figure S1. Observed means of cardiovascular phenotypes with significant monotonic temporal trends.**

Note. Bar plots show the mean value of each cardiovascular imaging or electrocardiographic phenotype across follow-up intervals after definitive chemoradiation therapy. The mean represents the observed proportion of evaluable patients with the new or worsened phenotype during each interval. Phenotypes shown were those with significant monotonic temporal trends in multivariable GEE models adjusted for age, sex, total radiation dose, and number of radiation fractions. Bar heights represent observed interval-specific means. Insets show adjusted odds ratios, 95% CIs, and *P* values from the multivariable generalized estimating equation models; odds ratios represent the average change in odds per one-category increase in follow-up interval.

Abbreviations: CI = confidence interval; GEE = generalized estimating equation; OR = odds ratio; PA = pulmonary artery.

| Imaging Variable | 0–6 mo | 6–12 mo | 1–2 yr | 2–5 yr | >5 yr |
|---|---|---|---|---|---|
| **Pericardial effusion** | I: 0.866<br>NI: 0.938<br>M: 0.080 | I: 0.205<br>NI: 0.683<br>M: 0.062 | I: 0.278<br>NI: 0.101<br>M: 0.503 | I: 0.178<br>**NI: 0.040***<br>M: 0.759 | I: 0.154†<br>NI: 0.176†<br>M: 1.000† |
| **Pericardial thickening** | I: 0.712<br>NI: 0.190<br>M: 0.250 | I: —<br>NI: —<br>M: — | I: 0.846<br>NI: **0.023***<br>M: 0.296 | I: —<br>NI: —<br>M: — | I: —<br>NI: —<br>M: 1.000† |
| **LV systolic dysfunction** | I: 1.000†<br>NI: 1.000†<br>M: 1.000† | I: 1.000†<br>NI: 1.000†<br>M: — | I: 1.000†<br>NI: 0.580†<br>M: 0.286† | I: 1.000†<br>NI: 1.000†<br>M: 1.000† | I: 0.333†<br>NI: 0.333†<br>M: — |
| **Regional wall motion abnormality** | I: 0.206<br>NI: 0.459<br>M: 0.261 | I: 1.000†<br>NI: 1.000†<br>M: 0.167† | I: 1.000†<br>NI: 1.000†<br>M: 1.000† | **I: 0.005***<br>**NI: 0.050***<br>**M: 0.013*** | I: 1.000†<br>NI: 1.000†<br>M: — |
| **Global hypokinesis** | I: 0.544<br>NI: 0.244<br>M: 0.544 | I: 0.400†<br>NI: 1.000†<br>M: — | I: 0.619<br>NI: 0.274<br>M: 1.000† | I: 0.629<br>NI: 0.375<br>M: 0.531 | I: 0.250†<br>NI: 0.250†<br>M: — |
| **Diastolic dysfunction** | I: 0.364<br>NI: 0.784<br>M: 0.484 | I: 0.152†<br>NI: 0.491†<br>M: 1.000† | I: 0.178<br>NI: 0.769<br>M: 1.000† | I: **0.004***<br>NI: 0.235<br>M: 0.394 | I: 1.000†<br>NI: 1.000†<br>M: — |
| **LV dilation** | I: 0.576<br>NI: 0.271<br>M: 0.536 | I: **0.045†***<br>NI: 0.091†<br>M: 1.000† | I: 0.732<br>NI: 0.502<br>M: 1.000† | I: 0.600<br>NI: 0.412<br>M: 0.555 | I: —<br>NI: —<br>M: — |
| **Cardiomegaly** | I: 0.639<br>NI: **0.028***<br>M: 0.185 | I: 0.644<br>NI: 0.587<br>M: 0.086 | I: 0.652<br>NI: 0.717<br>M: 0.136 | I: 0.698<br>NI: 0.552<br>M: 0.482 | I: 0.455†<br>NI: 0.491†<br>M: 1.000† |
| **Coronary calcification progression** | I: 0.795<br>NI: 0.643<br>M: — | I: 0.826<br>NI: 0.796<br>M: 0.306 | I: 0.849<br>NI: 0.662<br>M: 0.346 | I: 0.213<br>NI: 0.900<br>M: 0.457 | I: 0.525†<br>NI: 0.538†<br>M: 1.000† |

| | | | | | |
|---|---|---|---|---|---|
| **Perfusion abnormality** | I: —<br>NI: —<br>M: — | I: —<br>NI: —<br>M: — | I: 1.000†<br>NI: 1.000†<br>M: 1.000† | I: 1.000†<br>NI: 1.000†<br>M: 1.000† | I: —<br>NI: —<br>M: — |
| **Valvular calcification progression** | I: 0.796<br>NI: 0.646<br>M: — | I: —<br>NI: —<br>M: — | I: 0.849<br>NI: 0.662<br>M: 0.280 | I: 0.569<br>NI: **0.048***<br>M: 0.500 | I: 0.081†<br>NI: 0.093†<br>M: 1.000† |
| **Valvular dysfunction** | I: 0.052<br>NI: 0.279<br>M: 0.482 | I: 0.429†<br>NI: 1.000†<br>M: — | I: 0.396†<br>NI: 1.000†<br>M: 1.000† | I: 0.294†<br>NI: 0.153†<br>M: 0.620† | I: 1.000†<br>NI: 1.000†<br>M: — |
| **Thoracic aortic calcification progression** | I: —<br>NI: —<br>M: — | I: 0.826<br>NI: 0.796<br>M: 0.321 | I: **<.001***<br>NI: 0.183<br>M: 0.187 | I: 0.125<br>NI: 0.459<br>M: 0.147 | I: 0.569†<br>NI: 0.538†<br>M: 1.000† |
| **Main pulmonary artery enlargement** | I: 0.475<br>NI: 0.065<br>M: 0.807 | I: 0.541<br>NI: 0.473<br>M: 0.414 | I: 0.533<br>NI: 0.564<br>M: 0.358 | I: 0.386<br>NI: 0.635<br>M: 0.941 | I: 0.516†<br>NI: 1.000†<br>M: 0.250† |
| **Pulmonary hypertension** | I: 0.329<br>NI: 0.178<br>M: 0.456 | I: 0.143†<br>NI: 0.286†<br>M: — | I: —<br>NI: —<br>M: — | I: 1.000†<br>NI: 1.000†<br>M: 0.538† | I: 1.000†<br>NI: 1.000†<br>M: — |
| **Right heart strain** | I: 0.742<br>NI: 0.622<br>M: 0.444 | I: 0.073<br>NI: **0.002***<br>M: 0.560 | I: 0.535<br>NI: 0.301<br>M: 0.462 | I: 0.895<br>NI: 0.895<br>M: 0.944 | I: 1.000†<br>NI: 1.000†<br>M: 1.000† |
| **Conduction abnormality** | I: 0.417<br>**NI: 0.003***<br>M: 0.187 | I: —<br>NI: —<br>M: — | I: 0.529<br>**NI: <.001***<br>M: 0.221 | I: 0.389<br>**NI: <.001***<br>M: 0.197 | I: 1.000†<br>NI: 1.000†<br>M: — |
| **Arrhythmia** | I: 0.297<br>**NI: <.001***<br>M: 0.925 | I: 1.000†<br>NI: 0.231†<br>M: 1.000† | I: 0.364<br>**NI: 0.007***<br>M: 0.097 | I: 0.052<br>NI: 0.057<br>M: 0.752 | I: 1.000†<br>NI: 1.000†<br>M: — |

**Supplemental Table S4. Interval-specific associations between longitudinal cardiovascular phenotypes and clinical outcomes following definitive chemoradiation for lung cancer**

Note. Table shows interval-specific *P* values for associations between longitudinal cardiovascular imaging and electrocardiographic phenotypes and ischemic CVAEs, nonischemic CVAEs, and all-cause mortality.

*P ≤ 0.05 indicates statistical significance.

†Two-sided Fisher exact test was used because at least one expected cell count was less than 5. All other *P* values were obtained using Pearson $\chi^2$ tests.

— indicates that the association was not estimable because the imaging variable, electrocardiographic variable, or outcome lacked variation.

Abbreviations: CVAE = cardiovascular adverse event; I = ischemic CVAE; M = all-cause mortality; NI = nonischemic CVAE.

| Phenotype | Paired evaluable n | Exact agreement, % | Cohen κ | 95% CI | Interpretation/Note |
|---|---|---|---|---|---|
| **Cardiomegaly** | 15 | 100 | — | — | κ not estimable; no category variation |
| **Right-heart strain** | 15 | 93.3 | 0 | 0.000–0.000 | Sparse positives; single discordant positive rating |
| **Myocardial thinning** | 15 | 100 | — | — | κ not estimable; no category variation |
| **Myocardial attenuation** | 15 | 93.3 | 0 | 0.000–0.000 | Sparse positives; single discordant positive rating |
| **Pericardial thickening** | 15 | 100 | — | — | κ not estimable; no category variation |
| **Pericardial effusion** | 15 | 86.7 | 0.722 | 0.331–1.000 | Substantial agreement |
| **Coronary arterial calcification** | 15 | 100 | 1 | 1.000–1.000 | Perfect agreement |
| **Valvular calcification** | 15 | 93.3 | 0.762 | 0.283–1.000 | Substantial agreement |
| **Thoracic aortic calcification** | 15 | 100 | 1 | 1.000–1.000 | Perfect agreement |
| **Pulmonary artery enlargement** | 15 | 86.7 | 0.444 | −0.208–1.000 | Moderate agreement; limited by sparse positives |

**Supplemental Table S5. Interobserver agreement for binary cardiovascular imaging phenotype classifications**

Note. Phenotypes represent binary follow-up classifications of new or worsened abnormalities compared with baseline. Absent, unchanged, or improved findings were classified as negative. Agreement was assessed in a 15-case subset independently reviewed by two readers using unweighted Cohen κ with 95% confidence intervals. Exact agreement was also reported. κ was not estimable for variables with no category variation because both readers assigned all cases to the same category. Cohen κ values were interpreted according to the Landis and Koch classification: slight agreement, 0.00–0.20; fair, 0.21–0.40; moderate, 0.41–0.60; substantial, 0.61–0.80; and almost perfect, 0.81–1.00.

Abbreviations: CI = confidence interval.
